# High-fidelity long-read sequencing reveals a complex RCCX locus at the single-nucleotide level in Korean patients with congenital adrenal hyperplasia

**DOI:** 10.1101/2025.07.09.25331238

**Authors:** Seoyeon Kim, Ji-Hee Yoon, Dohyung Kim, Soyoung Park, Gu-Hwan Kim, Han-Wook Yoo, Jin-Ho Choi, Jun Kim, Ja Hye Kim

**Affiliations:** Graduate School of Life Sciences, College of Bioscience and Biotechnology, Chungnam National University, 99, Daehak-ro, Yuseong-gu, Daejeon 34134, Republic of Korea; Department of Pediatrics, Asan Medical Center, University of Ulsan College of Medicine, 88, Olympic-ro 43-gil, Songpa-gu, Seoul 05505, Republic of Korea; Asan Institute for Life Sciences, Asan Medical Center, University of Ulsan College of Medicine, 88, Olympic-ro 43-gil, Songpa-gu, Seoul 05505, Republic of Korea; Medical Genetics Center, Asan Medical Center, University of Ulsan College of Medicine, 88, Olympic-ro 43-gil, Songpa-gu, Seoul 05505, Republic of Korea; Department of Pediatrics, Bundang CHA Medical Center, CHA University, CHA University School of Medicine, Seongnam-si, 13496, Republic of Korea; Department of Convergent Bioscience and Informatics, College of Bioscience and Biotechnology, Chungnam National University, 99, Daehak-ro, Yuseong-gu, Daejeon 34134, Republic of Korea

**Keywords:** Long-read sequencing, Personalized genome assembly, Segmental duplication, Congenital adrenal hyperplasia, Structural variants

## Abstract

**Background:** The RCCX locus, resulting from segmental duplication, exhibits extensive sequence identity and modular variations because of unequal crossover events, leading to copy number variations and the formation of chimeric genes between active and pseudogenes. Precise characterization of this locus is essential for molecular diagnosis, because aberrations within this region can cause congenital adrenal hyperplasia (CAH), an autosomal recessive disorder. However, the intricate modular structures, comprising mono-, bi-, and tri-modular haplotypes, have posed challenges to accurate analysis using conventional sequencing technologies.

**Methods:** In this study, we analyzed the segmentally duplicated RCCX locus using a pangenome-based approach. High-fidelity long-read sequencing data were produced to generate high-quality personalized genome assemblies from patients with CAH. Their RCCX haplotypes were fully resolved at the single-nucleotide level, and compared with those from non-patient populations.

**Results:** We generated high-fidelity long-read sequencing data and constructed 26 phased genome assemblies from 13 Korean patients with CAH, achieving single-nucleotide resolution analysis of the RCCX locus. Furthermore, by integrating human draft pangenome reference data, we analyzed non-patient cohorts and identified characteristic variations unique to the Korean patient population and novel structural variants formed through DNA damage and homologous recombination.

**Conclusions:** Our findings provide a key methodological framework for resolving segmentally duplicated genomic regions associated with genetic disorders. This study is expected to serve as a foundation for the precise molecular diagnosis of patients with CAH.

**Graphical abstract:** 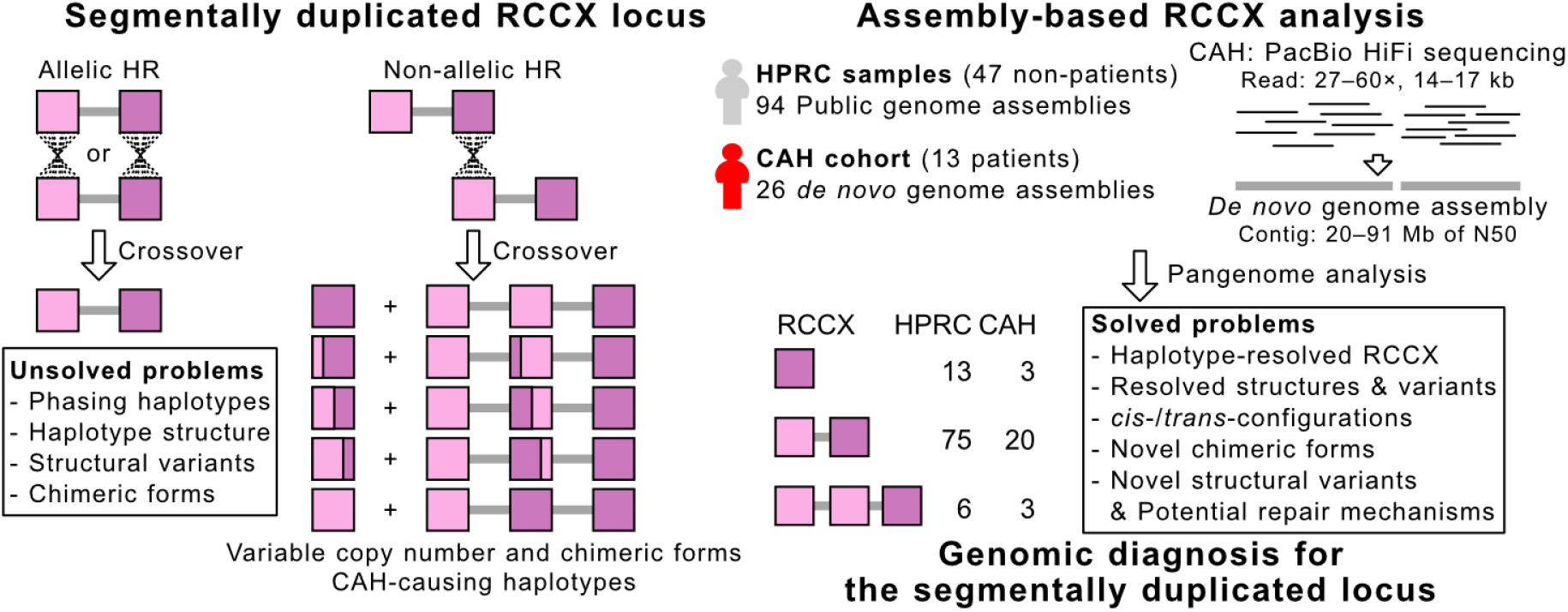

## Background

Congenital adrenal hyperplasia (CAH) is an autosomal recessive disorder characterized by enzyme deficiencies within the adrenal steroidogenic pathway, leading to impaired cortisol and aldosterone synthesis and consequent hormonal imbalances (1–3). Approximately 95% of CAH cases are caused by mutations in *CYP21A2*, which encodes steroid 21-hydroxylase, an enzyme critical for cortisol and aldosterone synthesis. These pathogenic variants impair the enzymatic activity of 21-hydroxylase, and the extent of residual enzymatic activity determines the clinical severity of CAH. CAH is classified into the classic type, comprising salt-wasting and simple virilizing forms, and the non-classics type (1, 4–6). Additionally, approximately 10% of patients with CAH are further classified as having CAH-X syndrome owing to contiguous gene deletions involving *CYP21A2* and *TNXB*. These patients exhibit clinical features of hypermobile Ehlers-Danlos syndrome (hEDS) because of tenascin-x deficiency, with biallelic *TNXB* mutations leading to more severe manifestations, including musculoskeletal abnormalities and cardiac valvulopathy (7–11).

*CYP21A2* is located on chromosome 6p21.3, within a segmentally duplicated region known as the RCCX locus (12). This locus is characterized by a modular structure that predisposes it to complex copy-number variations (CNVs) through mechanisms such as non-allelic homologous recombination (NAHR). Among individuals of Caucasian ancestry, bi-modular alleles account for 69.0–78.8%, whereas mono- and tri-modular alleles represent 14.1–17.0% and 7.1–14.0%, respectively (13–18). Individuals with four or five modules have rarely been reported (17, 19).

These CNVs are thought to arise from segmental duplications of the ancestral RCCX module, further compounded by unequal crossover events between functional genes and their pseudogene counterparts because of sequence homology (e.g., *STK19* and *STK19B*, *CYP21A2* and *CYP21A1P*, and *TNXB* and *TNXA*) (12, 20). Such recombination events are likely more frequent than *de novo* mutations specific to the *CYP21A2* gene, potentially contributing to the relatively higher incidence of CAH among rare genetic disorders (12, 15, 21). Detailed analysis of the RCCX locus, encompassing its modular structure and gene functionality, is critical for improving both the diagnosis and understanding of CAH. However, technical challenges have hampered such efforts because current genetic diagnostic methods are inadequate for resolving extensive and complex genomic regions. For example, multiplex ligation-dependent probe amplification (MLPA) combined with Sanger sequencing is commonly employed to detect genetic variants of *CYP21A2,* including CNVs. However, this approach cannot fully analyze the complex modular structure of the RCCX locus, particularly large-scale gene recombination events or the cis- or trans-configuration of multiple variants (12, 22–24). Although next-generation sequencing (NGS) methods have been utilized, their short-read lengths are insufficient for fully elucidating the highly repetitive and structurally complex nature of the RCCX locus (12, 25).

Recent advancements in long-read sequencing technologies have enabled analysis of extensive and complex genomic regions, including the RCCX locus (26–30). Platforms such as Pacific Biosciences (PacBio) generate 10–20 kb reads with a base-level accuracy of approximately 99.9% (31, 32). Several studies have employed these technologies to distinguish *CYP21A2* from its pseudogene, *CYP21A1P*, and to accurately identify pathogenic variants in *CYP21A2* or its chimeric forms by sequencing full-length *CYP21A2* of approximately 3 kb (33–37). However, read-level analyses remain insufficient to fully resolve the modular structures of the RCCX locus at the allele level, as this locus spans much longer sequences (approximately 105 kb for monomodular and 171 kb for trimodular alleles) than typical read lengths. Additionally, analysis of *TNXB* is particularly challenging owing to its extensive 68-kb genic region, which complicates accurate diagnosis in patients with CAH-X syndrome (33, 38).

These limitations can be overcome by using phased and more contiguous genomic sequences through the assembly of long reads (28, 39–47). With sufficient read depth, overlapping regions of long reads can be captured and assembled into longer sequences known as contigs (48–55). For instance, the Human Pangenome Reference Consortium (HPRC) recently published a draft human pangenome based on 94 phased genome assemblies from 47 healthy individuals (56, 57). Given that the average contig length of each phased genome assembly exceeds one million base pairs, these data can be leveraged to simultaneously identify large CNVs and small variants along with their allelic phase information. This approach allows the construction of two phased haploid sequences for each individual, offering a resolved representation of the RCCX locus without the complexity and ambiguity associated with reference-based read mapping. Despite these advantages, the modular structures of the RCCX locus in both the general population and patients with CAH remain poorly characterized, largely because of the limited availability of publicly accessible genome assemblies. Moreover, genome assemblies from individuals of Asian ancestry are scarce in public databases (28, 58, 59).

In this study, we generated high-fidelity long-read sequencing data from 13 Korean patients with CAH and made this dataset publicly available (Fig. 1). By comparing our data with the HPRC dataset, we aimed to identify variants specific to the Korean population and characterize genetic variations within the RCCX locus, including its modular structures. Specifically, we examined the distribution of modular types within the population, the combinations of active, pseudogene, and chimeric forms of *CYP21A2* and *CYP21A1P* or *TNXB* and *TNXA* for each modular type, and the pathogenic variants in our patients with CAH along with their allelic phase information. This is achievable only through assembly-level analysis, rather than read-level analysis, as it allows the exact resolution of the RCCX locus at full-length sequences. Additionally, we sought to identify any previously undetected structural variants within the RCCX locus. Our high-quality long-read sequencing and genome assembly resources will facilitate the investigation of genetic variants at the structural variant level within the Asian population and enable the precise diagnosis of CAH and CAH-X syndrome through phased, full-length characterization of the RCCX locus.

**Fig. 1.**
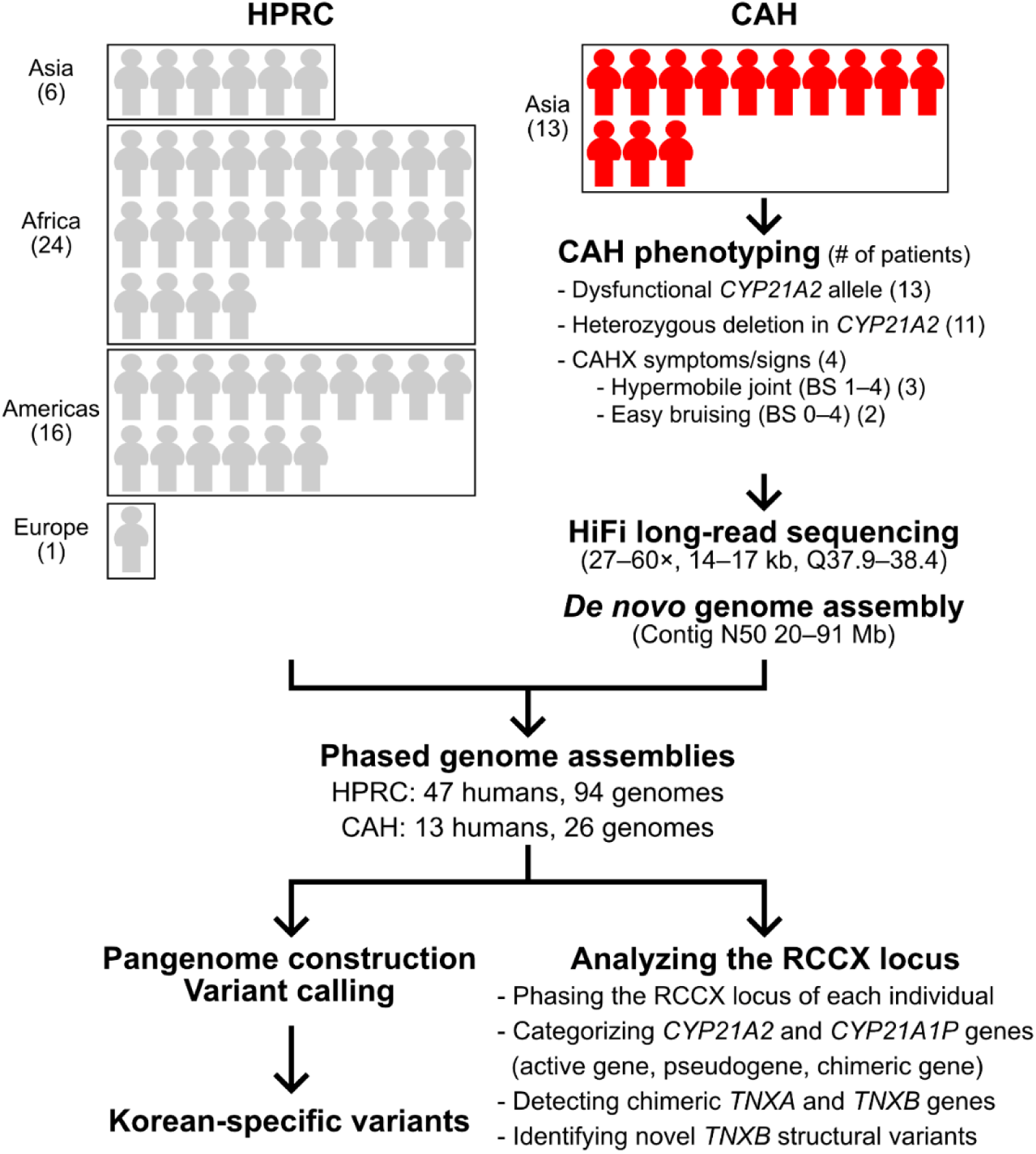
Experimental scheme for this study. Thirteen patients were diagnosed with CAH due to their dysfunctional *CYP21A2* alleles, which were identified using the MLPA–Sanger sequencing method. Of these, 11 patients were categorized as having at least one deletion allele, which is difficult to accurately analyze using conventional methods. HPRC genome assemblies were used to understand the RCCX structures of non-patients. These 120 genome assemblies from HPRC and our CAH patients were analyzed to identify Korean-specific variants and exact RCCX structures at the single-nucleotide level.

## Methods

### Cohort information and clinical phenotyping

Thirteen patients with CAH, including eight males and five females, were enrolled in this study. Clinical data were collected through a review of medical records from March 2023 to February 2024 at the Asan Medical Center, Seoul, Korea. The collected data included molecular analysis results confirmed by Sanger sequencing and MLPA analysis as well as the type of CAH. Additionally, all patients were evaluated for CAH-X syndrome based on their clinical symptoms and signs. Generalized joint hypermobility was determined using the Beighton score adjusted for age.

### DNA extraction and HiFi long-read sequencing

For DNA extraction, 5 mL of blood was collected from each patient. DNA extraction, sequencing library preparation, and HiFi long-read sequencing were performed by DNALink (https://dnalink.com/). Genomic DNA was extracted individually using a QIAGEN Genomic-tip 20/G. The extracted DNA was sheared into 15–18 kb fragments using the Megaruptor v3 system and purified with 1× SMRTbell cleanup beads. The sheared DNA underwent DNA repair, A-tailing, and adapter ligation, followed by a second purification step. Using the BluePippin system with the 0.75% DF Marker S1 High-Pass 6–10kb vs3 protocol, DNA fragments of specific lengths were selected, yielding the final PacBio HiFi SMRTbell sequencing libraries (15–20 kb). The libraries were sequenced using the PacBio Revio platform. The length and mean quality of the sequenced HiFi reads were determined using Bioawk (version 1.0; *bioawk -c fastx* with *length()* and *meanqual()* functions), and read length statistics were obtained using Assembly-stats (version 1.0.1; *assembly-stats*) (60).

### *De novo* genome assembly and pangenome construction

The HiFi long reads for each sample were assembled using Hifiasm (version 0.19.5-r587; default options) (48–50). To obtain contig length statistics for *de novo* genome assemblies, Assembly-stats was utilized (version 1.0.1; *assembly stats*) (60). Additionally, as previously described, the length statistics of the genome assemblies were processed to generate a cumulative contig length distribution (51). We performed the same contig length distribution analysis on the GRCh38.p14, T2T-CHM13v2.0, and HPRC (version v. 3. 0; *agc*) genome assemblies. The public genome assemblies used in this study were GRCh38.p14 (https://www.ncbi.nlm.nih.gov/datasets/genome/GCF_000001405.40/), T2T-CHM13v2.0 (https://www.ncbi.nlm.nih.gov/datasets/genome/GCF_009914755.1/), and the HPRC Year 1 assemblies (https://zenodo.org/record/5826274/files/HPRC-yr1.agc?download=1). A total of 120 genome assemblies obtained through this process were analyzed using the Minigraph-Cactus pipeline for pangenome analysis (version 2.6.7; *cactus-pangenome --giraffe clip filter - -vcf --viz --odgi --chrom-vg clip filter --chrom-og --gbz clip filter full --gfa clip full --vcf –giraffe --gfa --gbz --chrom-vg --logFile*) (61). The resulting VCF file from this analysis was subsequently used in our study.

### RCCX structure analysis

Sequences of *STK19* (chr6:31,971,175-31,981,446), *STK19B* (chr6:32,013,390-32,014,184), *CYP21A2* (chr6:32,038,415-32,041,644), *CYP21A1P* (chr6:32,005,636-32,008,909), *TNXA* (chr6:32,008,420-32,013,023), *TNXB* (chr6:32,041,153-32,109,338), *C4A* (chr6:31,982,057-32,002,681), *C4B* (chr6:32,014,795-32,035,418), and *HERV-K* (chr6:31,984,686-31,991,052) within the RCCX locus were extracted using SAMtools (version 1.18; *samtools faidx*), with genomic coordinates based on the NCBI RefSeq annotation of the GRCh38.p14 assembly (62).

To preliminarily determine the number of modules (RCCX CNVs), gene composition, and RCCX module structure for each genome assembly, we indexed all 120 genome assemblies and performed BLAST searches (version 2.14.0+; *makeblastdb -input_type fasta -dbtype nucl* and *blastn -task megablast -outfmt 6*) with the extracted genic sequences (*STK19*, *STK19B*, *HERV-K*, *C4A*, *C4B*, *CYP21A2*, *CYP21A1P*, *TNXB*, *TNXA*) (63). Because the active genes and pseudogenes of *STK* (*STK19* and *STK19B*) and *TNX* (*TNXB* and *TNXA*) differed in gene length, they were classified using both sequence identity and the ratio of the length of the aligned sequence to the actual gene length. Additionally, the presence of a known structural variant, a 120-bp deletion, in *TNXA* was considered in the classification of the *TNX* genes (8,12). For the *CYP* (*CYP21A2* and *CYP21A1P*) and *C4* (*C4A* and *C4B*) genes, where gene lengths were similar, sequence identity was the primary criterion for determination.

To achieve a more precise classification of the *CYP* and *TNX* gene types (active, pseudo, or chimeric genes), which are closely associated with CAH and CAH-X, we employed gene-specific variants. First, to identify markers distinguishing *CYP* gene types, we extracted all *CYP* genes with the 5’ upstream 5-kb regions from the 120 phased genome assemblies using SAMtools (version 1.18; *samtools faidx*) (62). These sequences were used as queries against the GRCh38 *CYP21A2* reference sequence using Minigraph-Cactus (version 2.6.7; *cactus-pangenome -- giraffe clip filter --vcf --viz --odgi --chrom-vg clip filter --chrom-og --gbz clip filter full --gfa clip full --vcf --giraffe --gfa --gbz --chrom-vg --logFile*), resulting in a VCF file containing variant information for *CYP* genes across all 120 genome assemblies (61). Variant annotation based on GRCh38 positions was performed using SnpEff (version 5.2a; GRCh38.p14) (64). Specifically, *CYP21A2* and *TNXB* variants were annotated using the NM_000500.9 and NM_001365276.2, respectively. We then selected pseudogene-specific variant markers to determine the *CYP* gene type. We first searched all *CYP* gene sequences identified from pangenome assemblies against the reference sequences, obtaining 111 *CYP21A2* active genes (48%), 115 *CYP21A1P* pseudogenes (49%), and 7 *CYP* chimeric genes (3%). By utilizing these allele frequencies a priori, we focused on variants with allele frequencies between 0.45 and 0.55 to identify pseudogene-specific markers. This is because variants resulting from pseudogenes, which occur in pseudogenes (49%) and chimeric genes (3%), are expected to have allele frequencies as high as 52% or as low as 46% in the population.

The variant marker data for each gene were subjected to dimensionality reduction using Principal Component Analysis (*PCA(n_components=3)*) via Scikit-learn, followed by clustering using the Agglomerative Clustering algorithm (*AgglomerativeClustering (n_clusters=4)*) to categorize all *CYP* genes into four types: *CYP21A2* active gene, *CYP21A1P* pseudogene, *CYP21A1P/CYP21A2* chimeric gene, and *CYP21A2/CYP21A1P* chimeric gene (65). A similar analysis was conducted for the *TNX* genes, using GRCh38 *TNXB* as the reference (focusing on variants within the region chr6:32,041,153–32,045,863 that exhibited allele frequencies between 0.45 and 0.60). Additionally, during this process, some variants were filtered out because they were present in both active genes and pseudogenes, and thus ineffective as markers.

By integrating all analyses, we determined the final haplotypic RCCX structure for each phased genome assembly. Furthermore, we analyzed the total variants in the *CYP21A2* and *TNXB* active genes for each genome assembly to identify the presence of known pathogenic variants associated with CAH or CAH-X (12, 66). Finally, by integrating our newly identified variant markers with markers commonly used in previous studies, we evaluated the subtypes of *CYP* and *TNX* chimeric genes and assessed their functional status (12).

## Results

### High-quality long-read data were obtained from 13 selected CAH patients

Thirteen Korean patients with congenital adrenal hyperplasia (CAH), confirmed to have pathogenic variants in *CYP21A2* by Sanger sequencing and/or multiplex ligation-dependent probe amplification (MLPA), were included in this study (Fig. 1). The median age at the time of data collection was 13 years (range, 5–42 years), and the cohort comprised 8 males and 5 females. Among the 13 patients, two (CAH-01 and CAH-02) carried only pathogenic single-nucleotide variants (SNVs), whereas the remaining 11 had at least one haplotype involving a large deletion of *CYP21A2* at the copy number variant (CNV) level. Four patients (CAH-06, CAH-10, CAH-12, and CAH-13) exhibited clinical features suggestive of CAH-X syndrome (Fig. 2A).

**Fig. 2.**
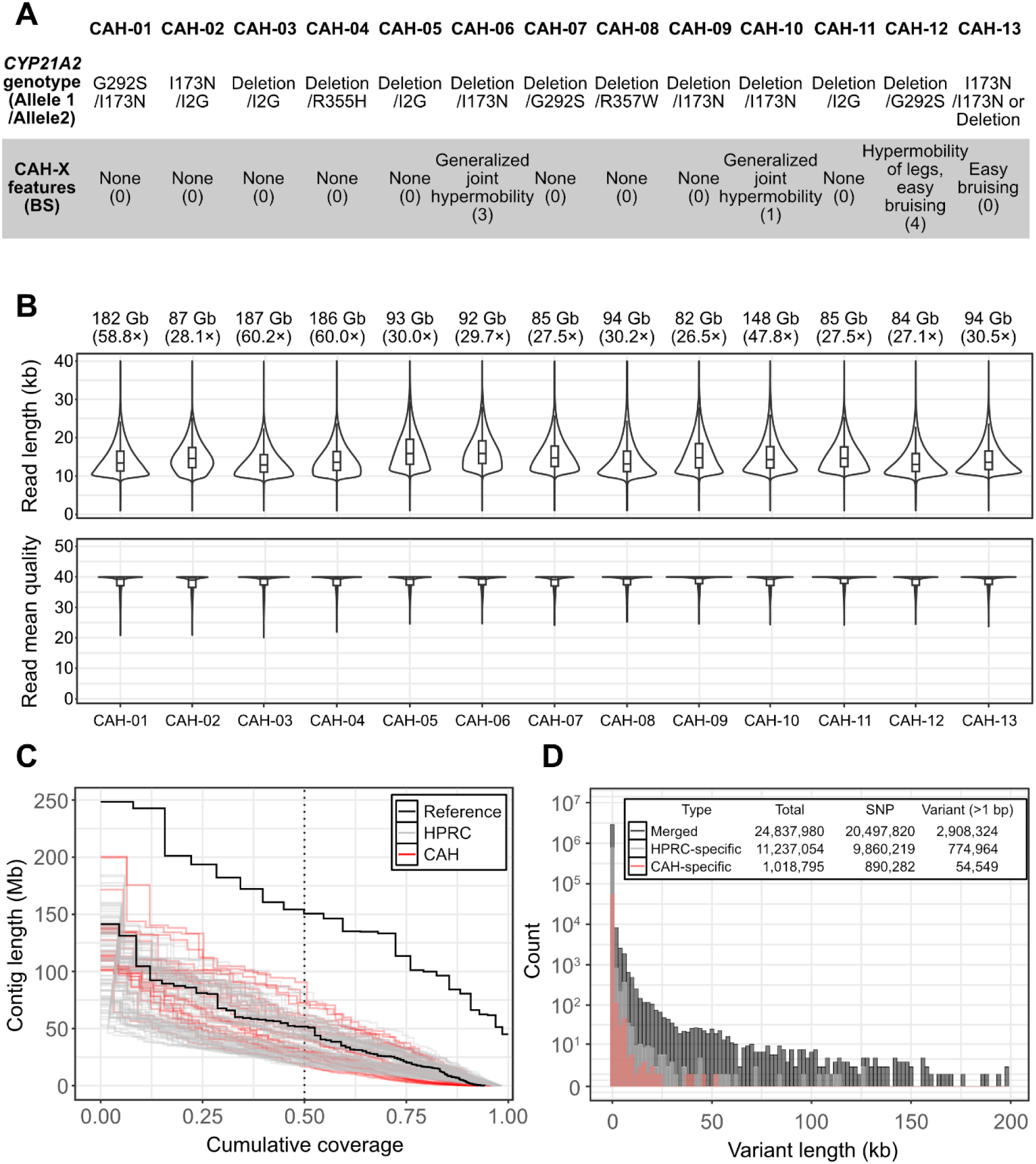
Patient information and quality assessment of HiFi long reads and genome assemblies. **A** Genotypes of *CYP21A2* and clinical features for CAH and CAH-X in 13 CAH patients. BS: Beighton score. **B** Length and quality score distribution of HiFi long-read sequencing data. At the top, the total amount of HiFi long-read sequencing data and sequencing depth for each sample are displayed. Horizontal lines within each violin plot represent the first quartile, median, and third quartile. **C** NGx plot of human genome assemblies. The x-axis shows the cumulative coverage of contig lengths relative to the T2T-CHM13 human genome size (3.1 Gb), whereas the y-axis displays the length of contigs. Contigs were sorted in descending order by length. The vertical dotted black line marks the NG50 value (20–91 Mb in CAH and 17–85 Mb in HPRC samples, respectively). The two reference lines correspond to T2T-CHM13 (upper line) and GRCh38 (lower line). **D** Length distribution of population-specific variants excluding single-nucleotide polymorphisms and single-nucleotide variants.

Using the PacBio Revio system, we obtained HiFi long-read sequencing data from 13 Korean patients with CAH. Each sample produced between 82 Gb and 187 Gb of HiFi reads, corresponding to approximately 27–60× read depths relative to the 3.1 Gb human genome (Fig. 2B). The HiFi reads showed a mean quality of Q37.88–Q38.40 (99.98–99.99% base-level accuracy) and an average read length of 14–17 kb, indicating both high accuracy and long read lengths (Table S1 and Fig. 2B).

### Nearly complete genome assemblies of CAH patients revealed potential Korean-specific variants

Using the HiFi data, we constructed two phased *de novo* genome assemblies per patient, each partially corresponding to paternally and maternally inherited haplotypes. Each draft genome assembly exceeded 2.9 Gb in size, suggesting near-complete genome coverage. The longest contigs ranged from 101 to 200 Mb, with N50 values of 20–91 Mb. Although these assemblies are not fully complete at the chromosome level, as achieved in the T2T-CHM13 assembly, they are comparable to or more contiguous than other high-quality reference genomes, including GRCh38 and 94 genome assemblies reported by the Human Pangenome Reference Consortium (HPRC) (52–184 Mb longest contigs; 17–85 Mb N50 values) (Table S2 and Fig. 2C) (56, 57).

Subsequently, we performed graph-based variant calling using a combined dataset of 26 Korean CAH patient assemblies, 94 HPRC assemblies, and the CHM13 genome, all referenced against GRCh38. Whereas 22 million variable loci had been identified in the previously reported HPRC data (94 assemblies), our expanded dataset revealed approximately 24.8 million variable loci (20,497,820 SNPs and 107,845 structural variants [SVs]), representing an increase of about 2.8 million loci (56, 57). Of these 24.8 million loci, 11.2 million were HPRC-specific (9.9 million SNPs, 18,395 SVs), whereas 1.0 million were specific to patients with CAH (890,282 SNPs, 1,809 SVs) (Fig. 2D). Notably, the CAH-specific variants included more than 50,000 small and large indels, with the longest reaching 52.8 kb. Considering that CAH results from dysfunctional *CYP21A2* and does not inherently predispose patients to accumulating additional *de novo* variants, these findings likely reflect variants specific to the Korean population, rather than being directly related to CAH itself (Fig. 2D). These Korean-specific variants were not observed on any sex chromosome, and no single chromosome exhibited variant enrichment; however, chromosome 19 displayed a relatively lower number of variants than the other autosomes (Fig. S1).

### Novel markers obtained from pangenome assemblies clarified exact haplotype structures in the RCCX locus

Using the phased genome assemblies of 47 unaffected individuals (from the Human Pangenome Reference Consortium [HPRC]) and 13 patients with CAH, a total of 60 individuals and 120 haplotype-level genome assemblies, we sought to characterize the copy number and gene composition in the RCCX region. To distinguish active genes from pseudogenes, we first surveyed all *CYP* and *TNX* sequences within the genome assemblies and identified marker variants that differentiated active genes from pseudogenes. From the 120 assemblies, we extracted 233 *CYP* and 238 *TNX* gene sequences. We identified genetic variants by comparing these gene sequences with the reference genome and determining allele frequencies. We selected 58 and 11 marker variants of *CYP* and *TNX*, respectively, based on their specificity to pseudogenes. These variants were present in 88–100% (99–112 of 112) of *CYP21A1P* alleles and 95–100% (101–106 of 106) of *TNXA* alleles, but only in 0–11% (0–12 of 105) of *CYP21A2* alleles and 0–8% (0–9 of 106) of *TNXB* alleles. These variants spanned a substantial portion of the genic regions of both genes: from the promoter to exon 10 of *CYP21A2* and from intron 34 to exon 43 of *TNXB*. Among these marker variants, we identified ten nonsynonymous variants in *CYP21A2* (p.P31L, p.G111Vfs, p.I173N, p.D184E, exon 6 cluster—p.I237N, p.V238E, p.M240K—, p.I237K, p.V238del, p.L308Ffs) and two in *TNXB* (120-bp deletion, p.Q3806R) (Tables S3 and S4).

Our marker set did not include all previously known variants used to differentiate the active, pseudo-, and chimeric forms of *CYP* and *TNX* genes. For example, two known *CYP21A1P*-specific variants, p.Q319X and p.R357W, were detected in only 74% (83/112) and 37% (41/112) of the *CYP21A1P* alleles, respectively (Table S5). Similarly, four known *TNXA*-specific markers, p.S4177N (85%; 90/106), p.D4174N (82%; 87/106), p.R4075H (91%; 96/106), and p.C4060W (68%; 72/106), were prevalent but not consistently observed across all *TNXA* alleles (Table S6). Therefore, these variants were excluded from our final marker set based on our stringent inclusion criteria, which required a high allele specificity. Using our final marker set, we classified 233 *CYP* sequences into 105 *CYP21A2* active genes (45%), 112 *CYP21A1P* pseudogenes (48%), 8 *CYP21A2/CYP21A1P* (3%), and 8 *CYP21A1P/CYP21A2* chimeric genes (3%). Similarly, 238 *TNX* genes were classified into 106 *TNXB* active genes (45%), 106 *TNXA* pseudogenes (45%), 4 *TNXB/TNXA* chimeric genes (2%), 14 *TNXA/TNXB* chimeric genes (6%), and 8 *TNXB* fragments (3%). We subsequently combined this gene-level information to determine the final gene composition of each RCCX haplotype across the 120 phased genome assemblies (Tables S3 and S4).

Analysis of the 120 haplotypes revealed that the majority contained *bimodular* RCCX structures with common homozygosity for the same module configuration. Specifically, 13% (16/120) were monomodular, 79% (95/120) were bimodular, and 8% (9/120) were trimodular (Fig. 3A). These frequencies are consistent with previous reports in *Caucasian* populations, where bimodular configuration (69–78.8%) is the most prevalent. Monomodular haplotypes were observed approximately 1.7 times more frequently than trimodular haplotypes, aligning with prior findings (1.2–2.0-fold difference) (13, 14, 17, 18). Moreover, 41 of the 60 individuals (68%) were homozygous at the module level (39 homozygous for bimodular and 2 for monomodular), whereas 19 individuals (32%) were heterozygous (10 carried mono-/bimodular, 7 carried tri-/bimodular, and 2 carried mono-/trimodular combinations) (Fig. S2).

**Fig. 3.**
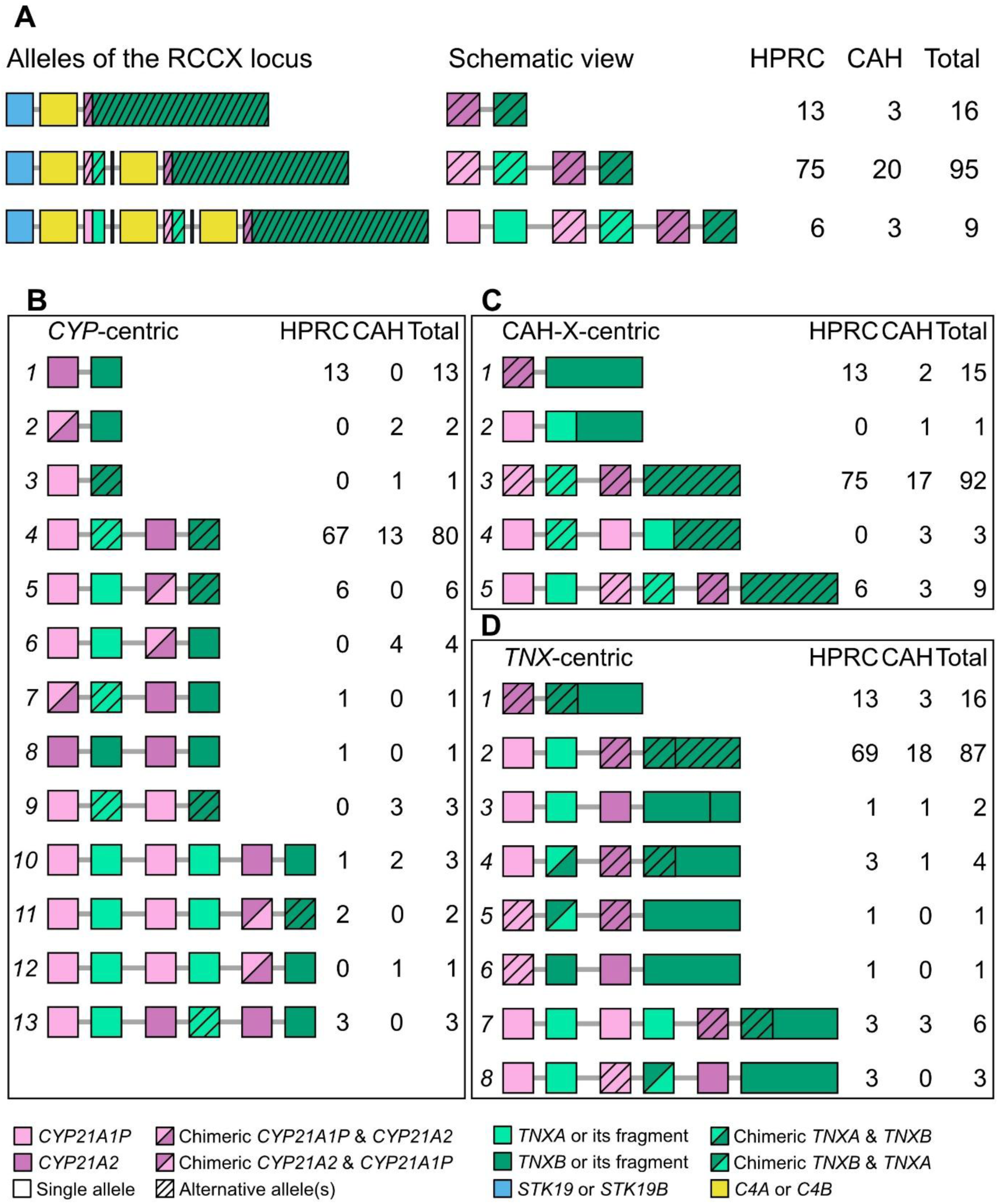
Population-level distribution of modular structures in the RCCX locus across our CAH (patient) and HPRC (non-patient) genome assemblies. **A** Overall distribution of RCCX module numbers. The left panel shows all genes within the RCCX locus, whereas the right panel represents schematic structures including only *CYP* and *TNX* genes. The other panels **B**–**D** also use this schematic representation. Although *CYP* and *TNX* genes overlap in the RCCX locus, they are displayed separately for clarity. **B** Distribution of RCCX haplotypes based on the type and number of *CYP* genes (*CYP21A2* active gene, *CYP21A1P* pseudogene, and their chimeric gene). Genotypes of *TNX* genes were simplified. **C** Distribution of RCCX haplotypes associated with the concurrent absence of functional *CYP21A2* and *TNXB* genes. The other genotypes were also visualized simply. **D** Distribution of RCCX haplotypes based on the type and number of *TNX* genes (*TNXB* active gene, *TNXA* pseudogene, their chimeric gene, and *TNXB* with structural variation). Genotypes of *CYP* genes were simplified. For panels **A**–**D**, genes that are not the primary focus of the respective criteria are represented in colors corresponding to their wild-type forms with or without hatching. If the actual gene at a given position differs from the wild-type (e.g., replaced by a pseudogene or chimeric form), the position is marked with hatching.

### *CYP*-centric analysis revealed detailed haplotype frequencies in patient and non-patient populations

Based on the type of *CYP* gene present, we identified 13 distinct types of RCCX haplotypes, comprising 6 bimodular, 3 monomodular, and 4 trimodular configurations. Among the bimodular types, the most frequent haplotype (80/120) consisted of one *CYP21A1P* pseudogene and one functional *CYP21A2* gene, representing the most common overall arrangement (Fig. 3B, type *4*). This haplotype was also most frequently observed in both the non-patient and patient groups. In addition, we identified another bimodular configuration carrying one *CYP21A1P* pseudogene and one *CYP* chimeric gene. The *CYP21A2/CYP21A1P* chimeric form was observed exclusively in non-patients, whereas the *CYP21A1P/CYP21A2* chimeric form was solely detected in patients (Fig. 3B, types *5* and *6*). All identified *CYP21A2/CYP21A1P* chimeric forms corresponded to the reciprocal form of the CH8 subtype, which retains functionality but harbors five *CYP21A1P*-derived variants in the 3′ UTR (c.*390A>G, c.*440C>T, c.*443T>C, c.*464T>C, c.*474C>T) (Table S7, HG01243h1, HG02080h2, HG02486h1, HG02630h1, HG03486h2, HG03516h2). Furthermore, among bimodular haplotypes in non-patients, we identified two types, in which the *CYP21A1P* pseudogene position was replaced by either a *CYP21A1P/CYP21A2* chimeric gene or an active *CYP21A2* gene, alongside a functional *CYP21A2* gene (Fig. 3B, types *7*–*8*). Lastly, bimodular haplotypes containing only pseudogenes (two *CYP21A1P* copies) were observed exclusively in 3 patients with CAH (Fig. 3B, type *9*).

Among the monomodular haplotypes, one carried a single functional *CYP21A2* gene and exhibited the second-highest allele frequency (13/120) among the 13 distinct RCCX haplotypes. Notably, this haplotype was observed only in non-patients (Fig. 3B, type *1*). In contrast, monomodular haplotypes lacking a functional gene, containing either a *CYP21A1P* pseudogene or *CYP21A1P/CYP21A2* chimeric gene, were identified exclusively in patients with CAH (Fig. 3B, types *2* and *3*).

In the trimodular haplotypes, two types were most frequently observed (three occurrences each): one carrying two pseudogenes and one active gene, and another carrying one pseudogene and two active genes (Fig. 3B, types *10* and *13*). The former was found in both the patient and non-patient groups, whereas the latter was detected only in the non-patient group. In addition, two trimodular types containing two pseudogenes and one chimeric gene were identified (Fig. 3B, types *11* and *12*). Similar to the bimodular type, only *CYP21A2/CYP21A1P* chimeric but functional genes were observed in the non-patient group. In contrast, only the *CYP21A1P/CYP21A2* chimeric gene was detected in the patient group, corresponding to a dysfunctional chimeric form (Table S7, HG02622h1, NA19240h2, and CAH-06h1).

### *TNX*-centric analysis revealed haplotypes potentially causing CAH-X and novel structural variants in *TNXB*

We identified one monomodular haplotype (one case) and one bimodular haplotype (three cases), both lacking functional *CYP21A2* and *TNXB* genes, exclusively in patients with CAH (Fig. 3C, types *2* and *4*). In these haplotypes, the *CYP21A1P* pseudogene and *TNXA/TNXB* chimeric gene were detected at the loci typically occupied by functional *CYP21A2* and *TNXB* active genes (Fig. 3C).

The 120 haplotypes were also classified based on the *TNX* gene types, resulting in the identification of eight distinct haplotypes (Fig. 3D and Fig. 4A). Only *TNXB* or *TNXA/TNXB* chimeric genes were detected at the *TNXB* active gene position. In contrast, the *TNXA* pseudogene position exhibited a broader diversity of *TNX* gene types, with gene lengths similar to or slightly longer than *TNXA* (4.60–4.98 kb compared to 4.60 kb). Among the bimodular haplotypes, we identified four *TNXA/TNXB* chimeric forms: two corresponding to a novel CH-2 subtype (Table S8, HG02717h1 and HG03098h1, both in the non-patient group) and two corresponding to the CH-2 subtype (CAH-07 and HG01928h2 from a non-patient) (Fig. 3D, type *4*, Fig. 4B, and Table S8). One *TNXB/TNXA* chimeric gene was exclusively detected in the non-patient group. This gene represents a new subtype that differs from those previously reported, and is presumably formed by crossover events between exons 41 and 43 (Fig. 3D, type *5*, Fig. 4B, and Table S8, HG02109h1). Moreover, an additional haplotype carrying a *TNXB* fragment instead of a chimeric gene was identified in one non-patient (HG03492h2) (Fig. 3D, type *6*). In trimodular haplotypes, *TNXB/TNXA* chimeric genes corresponding to the reciprocal form of the CH-2 subtype were found exclusively in three non-patient individuals (Fig. 3D, type *8*, Fig. 4B, and Table S8, HG01071h2, HG02717h2, and HG03540h1).

**Fig. 4.**
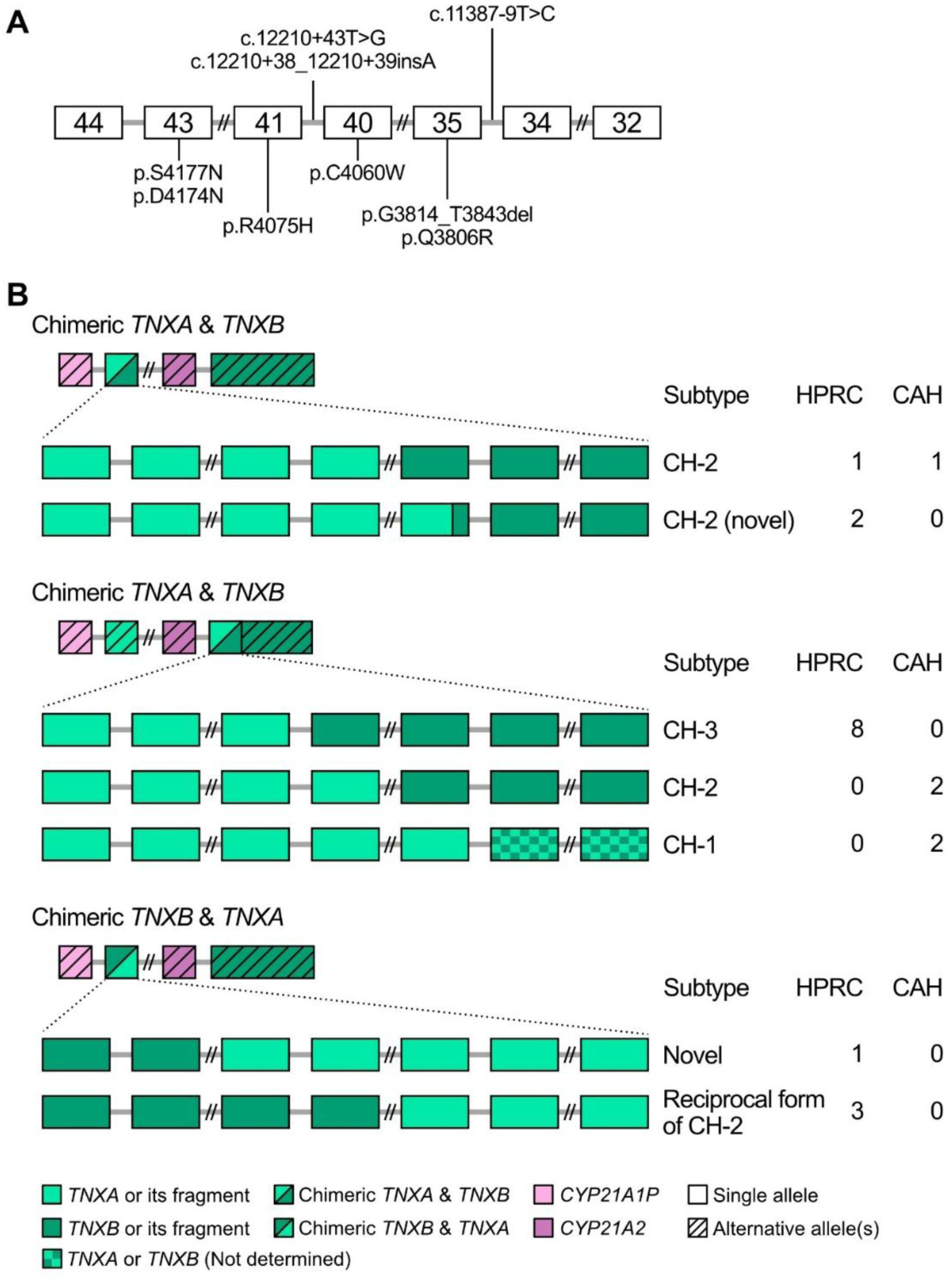
Chimeric configurations between *TNXA* and *TNXB* genes identified in the HPRC and CAH cohorts. **A** *TNXA*-specific variant markers used to identify *TNXA*- or *TNXB*-derived fragments. **B** Chimeric alleles are grouped by their position within the RCCX locus and by orientation (*TNXA* to *TNXB* or *TNXB* to *TNXA*); the subtype of each configuration and its allele counts are shown on the right.

Furthermore, we discovered two previously unreported structural variants within the 68-kb functional *TNXB* gene: a 3,339-bp deletion identified in one non-patient individual (NA18906h2) and a 68-bp insertion detected in two CAH patients (CAH-10 and CAH-12, Fig. 3C, type *4*, and Fig. 3D, types *2* and *3*). The 68-bp insertion represents tandem duplication of the intronic sequence at chr6:32,060,301–32,060,368 (intron 21 of *TNXB*). Notably, in addition to the 68-bp insertion, one haplotype also exhibited a *TNXA/TNXB* chimeric gene in the *TNXB* region (Fig. 3C, type *4*, and Fig. 4, CAH-12). The 3,339-bp deletion (chr6:32,058,155–32,061,493) spans intron 21 to exon 22, corresponding to c.7395_7727del, causing an in-frame deletion of 111 residues (p.Thr2465_Val2575del).

### Integrated haplotype and variant analyses elucidated the genetic basis of patient-specific *CYP21A2* and *TNXB* defects

Among the 26 haplotypes from patients with CAH, 15% (4/26) carried only pseudogenes, resulting in complete loss of *CYP21A2*. The remaining haplotypes carried either *CYP21A1P/A2* chimeric genes (27%, 7/26) or dysfunctional *CYP21A2* genes owing to other pathogenic variants (58%, 15/26) (Fig. 5).

**Fig. 5.**
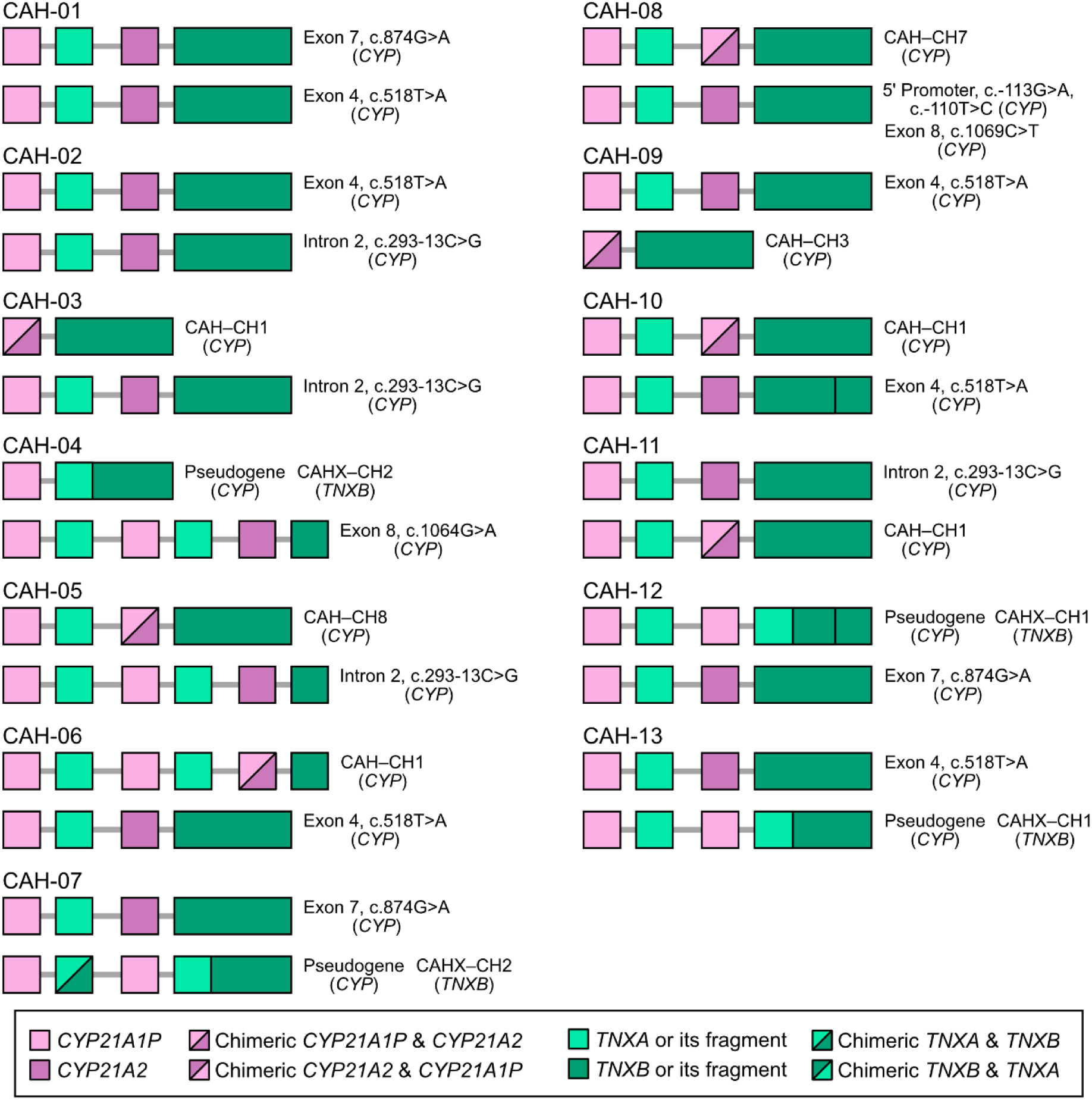
Haplotype structures of the RCCX region and pathogenic variant information of *CYP21A2* for each patient. Only the *CYP* and *TNX* genes within the RCCX locus are depicted, and the length of the trimodular *TNXB* is shown with the length omitted. To the right of each RCCX haplotype, the locations and genotypes of pathogenic variants in the *CYP21A2* gene, along with the resulting CAH phenotypes, are indicated. For haplotypes containing chimeric *TNXB* genes, the resulting CAH-X type is represented. Haplotypes containing only pseudogenes are labeled as “Pseudogene”. Chimeric *CYP* and *TNX* genes are annotated according to their chimeric types and junction sites.

We assessed whether *CYP21A1P/A2* chimeric genes or *CYP21A2* genes in these patient-derived haplotypes harbored pathogenic variants and classified chimeric subtypes. Compared to the reference *CYP21A2* sequence, the 15 patient-derived *CYP21A2* sequences carried 5–12 variants, encompassing six distinct pathogenic mutations (except CAH-02.h2, which had only 2 variants) (Table S9). In CAH-08, two promoter variants associated with reduced transcriptional activity and one pathogenic variant were identified in *cis* on the same haplotype, whereas all other *CYP21A2* alleles carried a single pathogenic variant associated with classic CAH phenotypes (Fig. 5 and Table S10) (67–69). Of the 10 reported *CYP21A1P/A2* chimeric gene subtypes, none of the alleles we observed in these patients were the attenuated forms (CH4, CH9, and CH10), which sometimes present milder phenotypes. Instead, we identified CH1 (4 alleles, CAH-03,06,10, and 11), CH3 (1 allele, CAH-09), CH7 (1 allele, CAH-08), and CH8 (1 allele, CAH-05). This aligns with previous reports that CH1 is the most frequently observed *CYP21A1P/A2* chimeric form (Fig. 5 and Table S7) (12, 33, 38, 70–73).

In patients with CAH lacking a functional *CYP21A2* gene, we further examined the integrity of the *TNXB* gene to identify cases likely to exhibit a CAH-X phenotype. This analysis revealed five CAH-X candidates, each harboring either a *TNXA/TNXB* chimeric gene or structural variants in *TNXB* that were predicted to impair its function (Fig. 5). Both CAH-12 and CAH-13 contain the CH-1-type *TNXA/TNXB* chimeric alleles (Fig. 4B and Fig. 5). Although these patients did not exhibit significant Beighton scores, their reported symptoms of hEDS suggest that the CH-1 chimeric allele may contribute to the CAH-X phenotype (Fig. 2A). In contrast, CAH-04 and CAH-07 had CH-2-type *TNXA/TNXB* chimeric alleles but did not exhibit clinical symptoms of hEDS (Fig. 4B). Unlike the aforementioned cases, CAH-10 did not harbor a *TNXA/TNXB* chimeric allele, but instead exhibited a 68-bp insertion in *TNXB*, a structural variant also observed in CAH-12, and presented with clinical features of hEDS. In addition to the five CAH-X candidates, CAH-06 presented with CAH-X symptoms; however, no previously reported pathogenic variant was detected in *TNXB* (Fig. 2A and Fig. 5).

## Discussion

Segmentally duplicated regions in the human genome pose major challenges to accurate variant detection and interpretation (27, 29, 30, 74). Among these, the RCCX locus is particularly complex because of its structural variability and the high sequence homology among its components (12, 15, 21). Precise identification of variants within this region, along with analysis of their population-level frequencies and associated functional consequences, is essential for understanding their clinical relevance (75–78). In this study, we used *de novo* genome assembly based on long-read sequencing to resolve the RCCX locus at single-nucleotide resolution. Unlike conventional approaches that primarily detect small or simple CNVs, our assembly-based strategy enabled the accurate differentiation of *CYP21A2* deletions arising from either pseudogenes or chimeric genes, as well as other coexisting variants and their *cis-* or *trans-*configurations (12, 22–24). This distinction provides deeper insight into the mechanisms by which each genetic configuration arises, thereby offering a more comprehensive understanding of CAH. Furthermore, this approach enabled the precise detection of genetic variants that distinguished patients with CAH from non-patient populations.

Compared to previous mapping-based methods, our assembly-based approach offers a significantly more robust strategy for analyzing segmentally duplicated regions. Previous studies have used locus-specific PCR followed by long-read sequencing to investigate RCCX-related genes, particularly *CYP21A2* and *TNXB*. Although these approaches have enabled the detection of known pathogenic variants, chimeric alleles, and gene rearrangements, they are inherently limited to predefined regions and depend on reference-based mapping (33–37, 79, 80). Consequently, they are unable to resolve the complete haplotype structure, detect complex CNVs, or provide phasing information across the entire RCCX locus. In contrast, our study applied *de novo* genome assembly based on long-read sequencing to comprehensively characterize the RCCX locus. This approach facilitated the reconstruction of the complete modular structure and gene content at the haplotype level. Additionally, it enabled the identification of previously unrecognized chimeric junctions and novel marker variants that could distinguish active genes from pseudogenes more reliably. By leveraging the full potential of long-read sequencing, our strategy overcomes the limitations of previous targeted analyses and establishes a new framework for high-resolution studies of segmentally duplicated loci.

Based on these advantages, we analyzed the RCCX locus—a highly variable, segmentally duplicated region—at the population level using human pangenome assemblies and confirmed that the frequencies of modular configurations were consistent with previous reports (13, 14, 17, 18, 81). Notably, the monomodular structure appeared at nearly twice the frequency of the trimodular structure in both the HPRC dataset and our CAH genome assemblies. This finding was unexpected, as monomodular and trimodular structures arise from bimodular structures via non-allelic homologous recombination (NAHR) and are therefore presumed to occur at comparable frequencies (15, 21). Another important observation was that while we identified a pathogenic monomodular configuration involving a chimeric *CYP21A1P*/*CYP21A2* gene, the pathogenic trimodular form (*CYP21A1P*– *CYP21A2*/*A1P*–*CYP21A2*) was not observed in any individual. These findings raise the possibility that monomodular structures may confer a phenotypic or evolutionary advantage over their trimodular counterparts. Further studies are needed to investigate whether such structural differences impact gene dosage, recombination dynamics, or selection pressures at the RCCX locus.

We successfully identified both the modules and the key genetic variants in the *CYP* and *TNX* genes within the RCCX locus. To the best of our knowledge, this is the first study to undertake a precise phased diagnosis of the RCCX locus in both non-patient and CAH patient populations. We identified five haplotypes in our CAH cohort in which *TNXB* was disrupted. Of these five haplotypes, four appeared to have resulted from NAHR between the *TNXA*-containing and *TNXB*-containing segments of homologous chromosomes (8, 82). In such cases, as observed in the CAH patients in this study, all *CYP* genes may be pseudogenes, and *TNXB* is rendered nonfunctional in the resulting haplotype. There are several plausible explanations for this structural pattern. First, the occurrence of NAHR between *TNXB* in one RCCX locus and *TNXA* in the other locus frequently results in the formation of a *TNXA/TNXB* chimera, in addition to the replacement of *CYP21A2* with the pseudogene *CYP21A1P*. This mechanism inherently links *TNXB* disruption together with the loss of a functional *CYP* gene. Second, such defective haplotypes may have arisen through recurrent NAHR events over generations, resulting in their stable inheritance in the population, even in the absence of strong selective pressure. However, it remains unclear whether these configurations are always inherited or arise *de novo*. To address this, trio-based analysis or parental genome assemblies are needed to assess the inheritance patterns and mutational origins of these complex haplotypes.

However, not all patients (CAH-04, CAH-07, CAH-12, and CAH-13) with these haplotypes displayed CAH-X features. CAH-04 and CAH-07, which harbor CH-2-type haplotypes, did not exhibit CAH-X features, unlike CAH-12 and CAH-13, which have the CH-1-type haplotype. Previous studies have reported that patients with biallelic CAH-X tend to manifest more severe clinical symptoms, including generalized joint hypermobility and skin hyperextensibility (38, 79). However, EDS-related traits were observed in a considerable proportion of monoallelic CAH-X individuals. In one study, approximately 58% of monoallelic patients exhibited at least one EDS feature, although the full clinical spectrum was not always present (79). Furthermore, the absence of symptoms in some younger patients suggests that age-dependent penetrance may contribute to the observed phenotypic variability (Fig. 2A and Fig. 5) (83).

To better understand the genetic factors that may underlie this variability in the clinical expression of CAH-X, we examined additional structural variants beyond canonical *TNXB* chimeras. We identified one haplotype that might disrupt *TNXB* through a previously unreported structural variant (a 68-bp insertion). This haplotype was observed in CAH-10 in the absence of the *TNX* chimeric gene. In addition, the same structural variant was also detected in CAH-12, which additionally carries a *TNX* chimeric gene and showed a higher Beighton score (BS) and clearer hEDS symptoms (Fig. 2A and Fig. 5). This variant could potentially affect the *TNXB* gene expression, but its clinical relevance remains uncertain. Notably, it remains to be determined whether such allele is also present in hEDS patients without CAH. Further studies, including functional assays and broader population screening, are necessary to clarify the pathogenic potential of this insertion. Questions remain regarding its penetrance, population frequency, and whether it can arise *de novo*. Accumulating additional genome assemblies from patients with CAH and their families is essential to address these issues.

Beyond known pathogenic variants and chimeric alleles, our high-resolution haplotype-level assemblies revealed additional layers of structural complexity within the RCCX locus. These included previously unrecognized structural variants and chimeric forms involving both *CYP* and *TNX*, providing new insights into the mutational mechanisms shaping this highly dynamic region. Although newly identified chimeric forms of *CYP* genes do not alter protein-coding sequences and are therefore unlikely to disrupt gene function, their presence expands the known mutational spectrum of this region. In *TNXB*, we identified not only the aforementioned 68-bp insertion but also a 3,339-bp deletion. In the case of the 3,339-bp deletion, we observed homologous flanking sequences with ∼330-bp stretch of homology. Such homology suggests the involvement of homology-directed repair (HDR) following a DNA double-strand break. More specifically, the relatively long stretches of homology are characteristic of single-strand annealing (SSA), an HDR mechanism distinct from polymerase theta–mediated end joining (TMEJ), which generally requires less than 20–30 bp of microhomology (28, 42, 47, 84) (Fig. 6A).

**Fig. 6.**
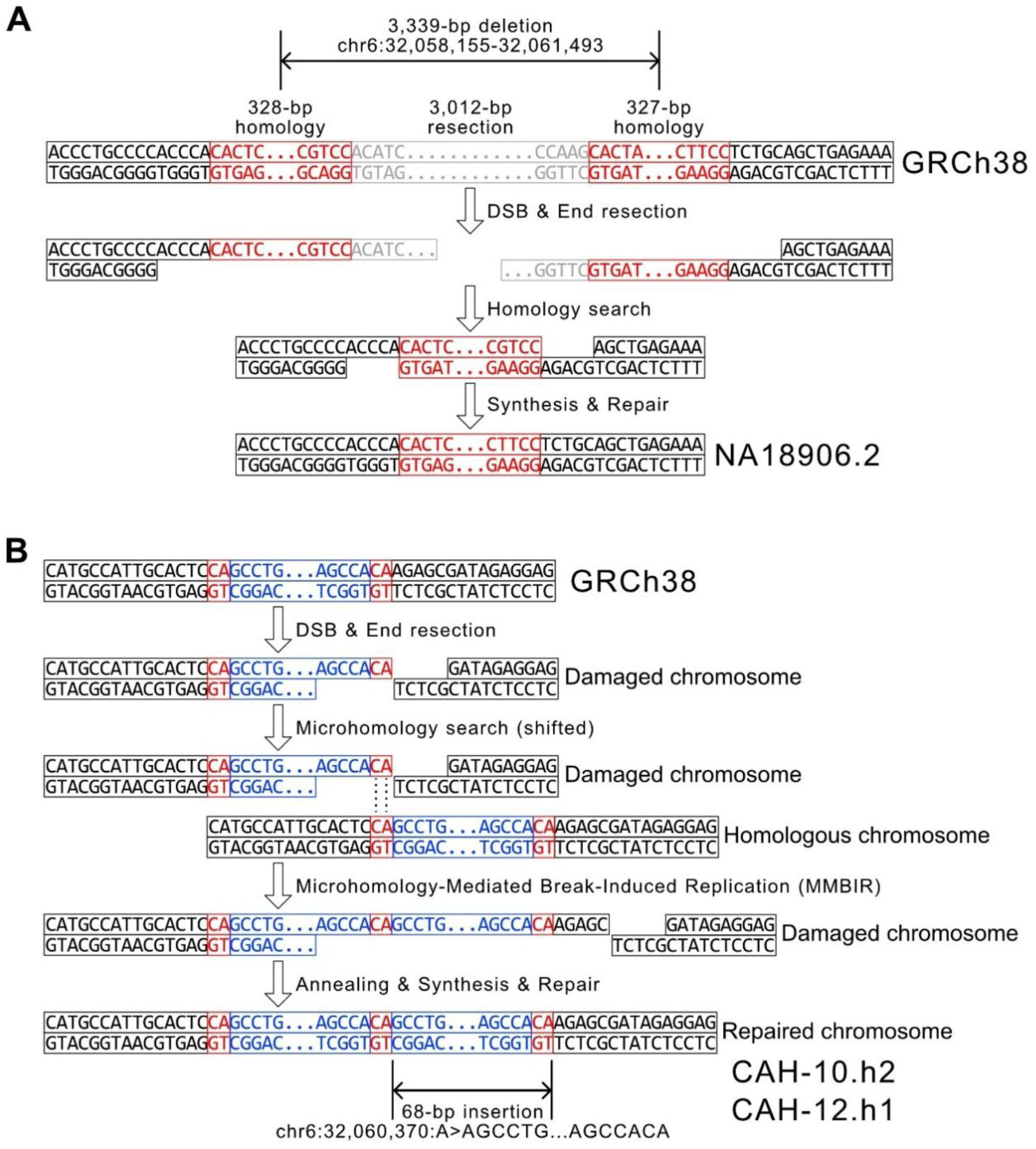
Possible DNA double-stranded break (DSB) repair mechanisms underlying the formation of novel structural variants. **A** Single-strand annealing (SSA) was visualized as the most plausible DSB repair mechanism for the 3,339-bp deletion, evidenced by its >300-bp homologous flanking sequences. **B** Microhomology-mediated break-induced replication (MMBIR) was visualized as the most plausible DSB repair mechanism for the 68-bp insertion, owing to its 2-bp microhomologous flanking sequences in addition to the duplicated sequence.

In the case of the 68-bp insertion, the direct duplication of an identical 68-bp sequence immediately adjacent to the insertion, flanked by 2-bp microhomologies at both ends, indicated that it most likely arose via microhomology-mediated break-induced replication (MMBIR), or a similar mechanism that repairs damaged ends by replicating a template sequence using microhomologous sequences (85, 86) (Fig. 6B). As the 68-bp insertion appeared in two individuals, these alleles may have originated from a shared ancestor. Meanwhile, the 3,339-bp deletion was observed only once in the HPRC data, and further investigation is needed to determine whether it affects *TNXB* function.

We generated high-quality genome assemblies from Korean individuals, thereby providing valuable resources for understanding the complex genetic variation within Asian populations. Although national pangenome projects are conducted globally, many remain incomplete, making it difficult to determine allele frequencies across different ancestral backgrounds (56, 58, 87–89). We believe that including data from rare disease cohorts can help bridge this gap. In particular, applying long-read sequencing to undiagnosed patients, whose conditions could not be fully elucidated by short-read sequencing, enables the identification of both causal variants and a broader range of population-specific genetic variation (90–99). To support such efforts, we have made the raw long-read sequencing data from 13 Korean individuals publicly available. As sequencing costs continue to decline and accuracy improves, we expect a rapid increase in the number of high-quality genome assemblies, enabling the detection of both common and rare variants, including complex and structurally challenging ones. Consequently, we anticipate that our genome assembly-based analyses will be widely adopted, ultimately establishing themselves as a standard methodology for the precise detection of a diverse range of complex genetic variants.

## Conclusions

Our study is the first to comprehensively elucidate previously elusive module structures, variant details, and *cis*-/*trans*-configuration information for the RCCX locus by leveraging state-of-the-art long-read sequencing technology and the resulting genome assemblies. By accumulating personalized genome assemblies, we can precisely capture both population-scale and patient-specific variant information. Moreover, this approach can be applied to other diseases caused by segmental duplications, such as spinal muscular atrophy (*SMN1*) and Gaucher’s disease (*GBA1*) (100, 101). We anticipate that ongoing innovations in sequencing technologies and computational tools will further advance the precision of genetic diagnostics.

## Data Availability

The raw sequencing data generated during the current study are available in the NCBI BioProject database, https://www.ncbi.nlm.nih.gov/bioproject/ , under the accession number PRJNA1233893.

## List of abbreviations

CAH: Congenital adrenal hyperplasia
hEDS: hypermobile Ehlers-Danlos syndrome
CNV: copy-number variations
NAHR: non-allelic homologous recombination
MLPA: multiplex ligation-dependent probe amplification
NGS: next-generation sequencing
PacBio: Pacific Biosciences
HPRC: Human Pangenome Reference Consortium
SNV: Single-nucleotide variant
BS: Beighton score
HDR: homology-directed repair
SSA: single-strand annealing
TMEJ: polymerase theta–mediated end joining
MMBIR: microhomology-mediated break-induced replication
DSB: DNA double-stranded break

## Declarations

### Ethics approval and consent to participate

This study was approved by the Institutional Review Board of Asan Medical Center (2023–0625).

### Consent for publication

Written informed consent for publication was obtained from all patients or their legal guardians. The consent permits the publication of relevant clinical information while explicitly prohibiting the disclosure of personally identifiable information.

### Availability of data and materials

The raw sequencing data generated during the current study are available in the NCBI BioProject database, https://www.ncbi.nlm.nih.gov/bioproject/ , under the accession number PRJNA1233893. The 94 pangenome assemblies of HPRC used during the current study are available in the Zenodo repository at https://zenodo.org/record/5826274/files/HPRC-yr1.agc?download=1 (HPRC-yr1.agc for HPRC Year 1 genome assemblies).

### Author’s contributions

Seoyeon Kim: Data curation, Formal analysis, Investigation, Methodology, Visualization, Writing—original draft. Ji-Hee Yoon: Data curation, Formal analysis, Investigation, Resources, Writing—review & editing. Dohyung Kim: Data curation, Formal analysis, Resources. Soyoung Park: Data curation, Formal analysis, Validation. Gu-Hwan Kim: Data curation, Formal analysis, Resources. Han-Wook Yoo: Data curation, Formal analysis, Resources. Jin-Ho Choi: Investigation, Methodology, Formal analysis, Data curation, Supervision, Conceptualization, Writing—review and editing. Jun Kim: Conceptualization, Funding acquisition, Project administration, Resources, Supervision, Visualization, Writing—original draft, Writing—review and editing. Ja Hye Kim: Conceptualization, Funding acquisition, Project administration, Resources, Supervision, Visualization, Validation, Writing—review and editing.

## Acknowledgements

We acknowledge and thank all the participating individuals.

## References

1. Claahsen-van der Grinten HL, Speiser PW, Ahmed SF, Arlt W, Auchus RJ, Falhammar H, et al. Congenital Adrenal Hyperplasia-Current Insights in Pathophysiology, Diagnostics, and Management. Endocr Rev. 2022;43(1):91–159.

2. Krone N, Arlt W. Genetics of congenital adrenal hyperplasia. Best Pract Res Clin Endocrinol Metab. 2009;23(2):181–92.

3. Speiser PW, White PC. Congenital adrenal hyperplasia. N Engl J Med. 2003;349(8):776–88.

4. Navarro-Zambrana AN, Sheets LR. Ethnic and National Differences in Congenital Adrenal Hyperplasia Incidence: A Systematic Review and Meta-Analysis. Horm Res Paediatr. 2023;96(3):249–58.

5. Pignatelli D, Carvalho BL, Palmeiro A, Barros A, Guerreiro SG, Macut D. The Complexities in Genotyping of Congenital Adrenal Hyperplasia: 21-Hydroxylase Deficiency. Front Endocrinol (Lausanne). 2019;10:432.

6. White PC, Speiser PW. Congenital adrenal hyperplasia due to 21-hydroxylase deficiency. Endocr Rev. 2000;21(3):245–91.

7. Lao Q, Brookner B, Merke DP. High-Throughput Screening for CYP21A1P-TNXA/TNXB Chimeric Genes Responsible for Ehlers-Danlos Syndrome in Patients with Congenital Adrenal Hyperplasia. J Mol Diagn. 2019;21(5):924–31.

8. Marino R, Moresco A, Perez Garrido N, Ramirez P, Belgorosky A. Congenital Adrenal Hyperplasia and Ehlers-Danlos Syndrome. Front Endocrinol (Lausanne). 2022;13:803226.

9. Merke DP, Chen W, Morissette R, Xu Z, Van Ryzin C, Sachdev V, et al. Tenascin-X haploinsufficiency associated with Ehlers-Danlos syndrome in patients with congenital adrenal hyperplasia. J Clin Endocrinol Metab. 2013;98(2):E379–87.

10. Miller WL, Merke DP. Tenascin-X, Congenital Adrenal Hyperplasia, and the CAH-X Syndrome. Horm Res Paediatr. 2018;89(5):352–61.

11. Morissette R, Chen W, Perritt AF, Dreiling JL, Arai AE, Sachdev V, et al. Broadening the Spectrum of Ehlers Danlos Syndrome in Patients With Congenital Adrenal Hyperplasia. J Clin Endocrinol Metab. 2015;100(8):E1143–52.

12. Kim JH, Kim GH, Yoo HW, Choi JH. Molecular basis and genetic testing strategies for diagnosing 21-hydroxylase deficiency, including CAH-X syndrome. Ann Pediatr Endocrinol Metab. 2023;28(2):77–86.

13. Bánlaki Z, Doleschall M, Rajczy K, Fust G, Szilágyi A. Fine-tuned characterization of RCCX copy number variants and their relationship with extended MHC haplotypes. Genes Immun. 2012;13(7):530–5.

14. Blanchong CA, Zhou B, Rupert KL, Chung EK, Jones KN, Sotos JF, et al. Deficiencies of human complement component C4A and C4B and heterozygosity in length variants of RP-C4-CYP21-TNX (RCCX) modules in caucasians. The load of RCCX genetic diversity on major histocompatibility complex-associated disease. J Exp Med. 2000;191(12):2183–96.

15. Carrozza C, Foca L, De Paolis E, Concolino P. Genes and Pseudogenes: Complexity of the RCCX Locus and Disease. Front Endocrinol (Lausanne). 2021;12:709758.

16. Gitelman SE, Bristow J, Miller WL. Mechanism and consequences of the duplication of the human C4/P450c21/gene X locus. Mol Cell Biol. 1992;12(5):2124–34.

17. Yang Y, Chung EK, Wu YL, Savelli SL, Nagaraja HN, Zhou B, et al. Gene copy-number variation and associated polymorphisms of complement component C4 in human systemic lupus erythematosus (SLE): low copy number is a risk factor for and high copy number is a protective factor against SLE susceptibility in European Americans. Am J Hum Genet. 2007;80(6):1037–54.

18. Yang Z, Mendoza AR, Welch TR, Zipf WB, Yu CY. Modular variations of the human major histocompatibility complex class III genes for serine/threonine kinase RP, complement component C4, steroid 21-hydroxylase CYP21, and tenascin TNX (the RCCX module). A mechanism for gene deletions and disease associations. J Biol Chem. 1999;274(17):12147–56.

19. Zhou D, Rudnicki M, Chua GT, Lawrance SK, Zhou B, Drew JL, et al. Human Complement C4B Allotypes and Deficiencies in Selected Cases With Autoimmune Diseases. Front Immunol. 2021;12:739430.

20. Demirdas S, Dulfer E, Robert L, Kempers M, van Beek D, Micha D, et al. Recognizing the tenascin-X deficient type of Ehlers-Danlos syndrome: a cross-sectional study in 17 patients. Clin Genet. 2017;91(3):411–25.

21. Bánlaki Z, Szabó JA, Szilágyi Á, Patócs A, Prohászka Z, Füst G, et al. Intraspecific evolution of human RCCX copy number variation traced by haplotypes of the CYP21A2 gene. Genome Biol Evol. 2013;5(1):98–112.

22. Baumgartner-Parzer S, Witsch-Baumgartner M, Hoeppner W. EMQN best practice guidelines for molecular genetic testing and reporting of 21-hydroxylase deficiency. Eur J Hum Genet. 2020;28(10):1341–67.

23. Concolino P, Mello E, Minucci A, Zuppi C, Capoluongo E. Multiplex ligation-dependent probe amplification analysis is useful for diagnosing congenital adrenal hyperplasia but requires a deep knowledge of CYP21A2 genetics. Clin Chem. 2011;57(7):1079–80.

24. Yoon JH, Hwang S, Kim JH, Kim GH, Yoo HW, Choi JH. Prenatal diagnosis of congenital adrenal hyperplasia due to 21-hydroxylase deficiency through molecular genetic analysis of the CYP21A2 gene. Ann Pediatr Endocrinol Metab. 2024;29(1):54–9.

25. Wang W, Han R, Yang Z, Zheng S, Li H, Wan Z, et al. Targeted gene panel sequencing for molecular diagnosis of congenital adrenal hyperplasia. J Steroid Biochem Mol Biol. 2021;211:105899.

26. Del Gobbo GF, Boycott KM. The additional diagnostic yield of long-read sequencing in undiagnosed rare diseases. Genome Research. 2025.

27. Jeong H, Dishuck PC, Yoo D, Harvey WT, Munson KM, Lewis AP, et al. Structural polymorphism and diversity of human segmental duplications. Nature Genetics. 2025;57(2):390–401.

28. Kim J, Park Jong L, Yang Jin O, Kim S, Joe S, Park G, et al. Highly accurate Korean draft genomes reveal structural variation highlighting human telomere evolution. Nucleic Acids Research. 2025;53(1).

29. Vollger MR, Dishuck PC, Harvey WT, DeWitt WS, Guitart X, Goldberg ME, et al. Increased mutation and gene conversion within human segmental duplications. Nature. 2023;617(7960):325-34.

30. Vollger MR, Guitart X, Dishuck PC, Mercuri L, Harvey WT, Gershman A, et al. Segmental duplications and their variation in a complete human genome. Science. 2022;376(6588):eabj6965.

31. Baid G, Cook DE, Shafin K, Yun T, Llinares-López F, Berthet Q, et al. DeepConsensus improves the accuracy of sequences with a gap-aware sequence transformer. Nature Biotechnology. 2023;41(2):232–8.

32. Wenger AM, Peluso P, Rowell WJ, Chang P-C, Hall RJ, Concepcion GT, et al. Accurate circular consensus long-read sequencing improves variant detection and assembly of a human genome. Nature Biotechnology. 2019;37(10):1155–62.

33. Adachi E, Nakagawa R, Tsuji-Hosokawa A, Gau M, Kirino S, Yogi A, et al. A MinION-based Long-Read Sequencing Application With One-Step PCR for the Genetic Diagnosis of 21-Hydroxylase Deficiency. J Clin Endocrinol Metab. 2024;109(3):750–60.

34. Li H, Zhu X, Yang Y, Wang W, Mao A, Li J, et al. Long-read sequencing: An effective method for genetic analysis of CYP21A2 variation in congenital adrenal hyperplasia. Clin Chim Acta. 2023;547:117419.

35. Liu Y, Chen M, Liu J, Mao A, Teng Y, Yan H, et al. Comprehensive Analysis of Congenital Adrenal Hyperplasia Using Long-Read Sequencing. Clin Chem. 2022;68(7):927–39.

36. Stephens Z, Milosevic D, Kipp B, Grebe S, Iyer RK, Kocher JA. PB-Motif-A Method for Identifying Gene/Pseudogene Rearrangements With Long Reads: An Application to CYP21A2 Genotyping. Front Genet. 2021;12:716586.

37. Zhang R, Cui D, Song C, Ma X, Cai N, Zhang Y, et al. Evaluating the efficacy of a long-read sequencing-based approach in the clinical diagnosis of neonatal congenital adrenocortical hyperplasia. Clin Chim Acta. 2024;555:117820.

38. Marino R, Garrido NP, Ramirez P, Notaristéfano G, Moresco A, Touzon MS, et al. Ehlers-Danlos Syndrome: Molecular and Clinical Characterization of TNXA/TNXB Chimeras in Congenital Adrenal Hyperplasia. J Clin Endocrinol Metab. 2021;106(7):e2789–e802.

39. Joe S, Park J-L, Kim J, Kim S, Park J-H, Yeo M-K, et al. Comparison of structural variant callers for massive whole-genome sequence data. BMC Genomics. 2024;25(1):318.

40. Kim C, Kim J, Kim S, Cook DE, Evans KS, Andersen EC, et al. Long-read sequencing reveals intra-species tolerance of substantial structural variations and new subtelomere formation in C. elegans. Genome research. 2019;29(6):1023–35.

41. Kim C, Sung S, Kim J, Lee J. Repair and reconstruction of telomeric and subtelomeric regions and genesis of new telomeres: implications for chromosome evolution. Bioessays. 2020;42(6):1900177.

42. Kim E, Kim J, Kim C, Lee J. Long-read sequencing and de novo genome assemblies reveal complex chromosome end structures caused by telomere dysfunction at the single nucleotide level. Nucleic Acids Research. 2021;49(6):3338–53.

43. Kim J, Kim Y, Shin J, Kim Y-K, Lee DH, Park J-W, et al. Fully phased genome assemblies and graph-based genetic variants of the olive flounder, Paralichthys olivaceus. Scientific Data. 2024;11(1):1193.

44. Kim J, Lim J, Kim M, Lee YK. Whole-genome sequencing of 13 Arctic plants and draft genomes of Oxyria digyna and Cochlearia groenlandica. Scientific Data. 2024;11(1):793.

45. Lee H, Kim J, Lee J. Benchmarking datasets for assembly-based variant calling using high-fidelity long reads. BMC Genomics. 2023;24(1):148.

46. Lim J, Kim W, Kim J, Lee J. Telomeric repeat evolution in the phylum Nematoda revealed by high-quality genome assemblies and subtelomere structures. Genome Research. 2023;33(11):1947–57.

47. Ryu H, Han H, Kim C, Kim J. GDBr: genomic signature interpretation tool for DNA double-strand break repair mechanisms. Nucleic Acids Research. 2025;53(2).

48. Cheng H, Asri M, Lucas J, Koren S, Li H. Scalable telomere-to-telomere assembly for diploid and polyploid genomes with double graph. Nature Methods. 2024;21(6):967–70.

49. Cheng H, Concepcion GT, Feng X, Zhang H, Li H. Haplotype-resolved de novo assembly using phased assembly graphs with hifiasm. Nature Methods. 2021;18(2):170–5.

50. Cheng H, Jarvis ED, Fedrigo O, Koepfli K-P, Urban L, Gemmell NJ, et al. Haplotype-resolved assembly of diploid genomes without parental data. Nature Biotechnology. 2022;40(9):1332–5.

51. Kim J, Kim C. A beginner’s guide to assembling a draft genome and analyzing structural variants with long-read sequencing technologies. STAR protocols. 2022;3(3):101506.

52. Koren S, Walenz BP, Berlin K, Miller JR, Bergman NH, Phillippy AM. Canu: scalable and accurate long-read assembly via adaptive k-mer weighting and repeat separation. Genome research. 2017;27(5):722–36.

53. Marx V. Method of the year: long-read sequencing. Nature Methods. 2023;20(1):6–11.

54. Nurk S, Walenz BP, Rhie A, Vollger MR, Logsdon GA, Grothe R, et al. HiCanu: accurate assembly of segmental duplications, satellites, and allelic variants from high-fidelity long reads. Genome research. 2020;30(9):1291–305.

55. Rautiainen M, Nurk S, Walenz BP, Logsdon GA, Porubsky D, Rhie A, et al. Telomere-to-telomere assembly of diploid chromosomes with Verkko. Nature Biotechnology. 2023;41(10):1474–82.

56. Liao W-W, Asri M, Ebler J, Doerr D, Haukness M, Hickey G, et al. A draft human pangenome reference. Nature. 2023;617(7960):312–24.

57. Wang T, Antonacci-Fulton L, Howe K, Lawson HA, Lucas JK, Phillippy AM, et al. The Human Pangenome Project: a global resource to map genomic diversity. Nature. 2022;604(7906):437-46.

58. Gao Y, Yang X, Chen H, Tan X, Yang Z, Deng L, et al. A pangenome reference of 36 Chinese populations. Nature. 2023;619(7968):112–21.

59. Nassir N, A. Almarri M, Akter H, Hassan Khansaheb H, Uddin KMF, Abou Tayoun A, et al. Advancing clinical genomics with Middle Eastern and South Asian pangenomes. Nature Medicine. 2025;31(3):725–7.

60. Trizna M. assembly_stats 0.1. 4. Zenodo. 2020.

61. Hickey G, Monlong J, Ebler J, Novak AM, Eizenga JM, Gao Y, et al. Pangenome graph construction from genome alignments with Minigraph-Cactus. Nat Biotechnol. 2024;42(4):663–73.

62. Danecek P, Bonfield JK, Liddle J, Marshall J, Ohan V, Pollard MO, et al. Twelve years of SAMtools and BCFtools. Gigascience. 2021;10(2).

63. Altschul SF, Gish W, Miller W, Myers EW, Lipman DJ. Basic local alignment search tool. J Mol Biol. 1990;215(3):403–10.

64. Cingolani P, Platts A, Wang le L, Coon M, Nguyen T, Wang L, et al. A program for annotating and predicting the effects of single nucleotide polymorphisms, SnpEff: SNPs in the genome of Drosophila melanogaster strain w1118; iso-2; iso-3. Fly (Austin). 2012;6(2):80–92.

65. Pedregosa F, Varoquaux G, Gramfort A, Michel V, Thirion B, Grisel O, et al. Scikit-learn: Machine learning in Python. the Journal of machine Learning research. 2011;12:2825–30.

66. Simonetti L, Bruque CD, Fernández CS, Benavides-Mori B, Delea M, Kolomenski JE, et al. CYP21A2 mutation update: Comprehensive analysis of databases and published genetic variants. Hum Mutat. 2018;39(1):5–22.

67. Araujo RS, Billerbeck AE, Madureira G, Mendonca BB, Bachega TA. Substitutions in the CYP21A2 promoter explain the simple-virilizing form of 21-hydroxylase deficiency in patients harbouring a P30L mutation. Clin Endocrinol (Oxf). 2005;62(2):132–6.

68. Zhang HJ, Yang J, Zhang MN, Zhang W, Liu JM, Wang WQ, et al. Variations in the promoter of CYP21A2 gene identified in a Chinese patient with simple virilizing form of 21-hydroxylase deficiency. Clin Endocrinol (Oxf). 2009;70(2):201–7.

69. Zhao Z, Gao Y, Lu L, Tong A, Chen S, Zhang W, et al. The underlying cause of the simple virilizing phenotype in patients with 21-hydroxylase deficiency harboring P31L variant. Front Endocrinol (Lausanne). 2022;13:1015773.

70. Chen W, Xu Z, Sullivan A, Finkielstain GP, Van Ryzin C, Merke DP, et al. Junction site analysis of chimeric CYP21A1P/CYP21A2 genes in 21-hydroxylase deficiency. Clin Chem. 2012;58(2):421–30.

71. Lan T, Wang J, Chen K, Zhang J, Chen X, Yao H. Comparison of long-read sequencing and MLPA combined with long-PCR sequencing of CYP21A2 mutations in patients with 21-OHD. Front Genet. 2024;15:1472516.

72. Lao Q, Burkardt DD, Kollender S, Faucz FR, Merke DP. Congenital adrenal hyperplasia due to two rare CYP21A2 variant alleles, including a novel attenuated CYP21A1P/CYP21A2 chimera. Mol Genet Genomic Med. 2023;11(7):e2195.

73. Zhang X, Gao Y, Lu L, Cao Y, Zhang W, Wu X, et al. Chimeric CYP21A1P/CYP21A2 Genes in 21-Hydroxylase Deficiency Detected by Long-Read Sequencing and Phenotypes Correlation. J Clin Endocrinol Metab. 2025.

74. Sharp AJ, Locke DP, McGrath SD, Cheng Z, Bailey JA, Vallente RU, et al. Segmental duplications and copy-number variation in the human genome. The American Journal of Human Genetics. 2005;77(1):78–88.

75. Auton A, Brooks LD, Durbin RM, Garrison EP, Kang HM, Korbel JO, et al. A global reference for human genetic variation. Nature. 2015;526(7571):68-74.

76. Karczewski KJ, Francioli LC, Tiao G, Cummings BB, Alföldi J, Wang Q, et al. The mutational constraint spectrum quantified from variation in 141,456 humans. Nature. 2020;581(7809):434-43.

77. MacArthur DG, Manolio TA, Dimmock DP, Rehm HL, Shendure J, Abecasis GR, et al. Guidelines for investigating causality of sequence variants in human disease. Nature. 2014;508(7497):469-76.

78. Richards S, Aziz N, Bale S, Bick D, Das S, Gastier-Foster J, et al. Standards and guidelines for the interpretation of sequence variants: a joint consensus recommendation of the American College of Medical Genetics and Genomics and the Association for Molecular Pathology. Genet Med. 2015;17(5):405–24.

79. Wang R, Luo X, Sun Y, Liang L, Mao A, Lu D, et al. Long-Read Sequencing Solves Complex Structure of CYP21A2 in a Large 21-Hydroxylase Deficiency Cohort. J Clin Endocrinol Metab. 2025;110(2):406–16.

80. Wang Y, Zhu G, Li D, Pan Y, Li R, Zhou T, et al. High clinical utility of long-read sequencing for precise diagnosis of congenital adrenal hyperplasia in 322 probands. Hum Genomics. 2025;19(1):3.

81. Liao WW, Asri M, Ebler J, Doerr D, Haukness M, Hickey G, et al. A draft human pangenome reference. Nature. 2023;617(7960):312–24.

82. Concolino P, Falhammar H. CAH-X Syndrome: Genetic and Clinical Profile. Mol Diagn Ther. 2022;26(3):293–300.

83. Figueras LM, Pacheco RM, González DG, Domènech MA, Zubicaray BE. Molecular characterization of the new clinical entity associated with congenital adrenal hyperplasia: the CAH-X syndrome in the Spanish population. Adv Lab Med. 2023;4(3):258–67.

84. Al-Zain AM, Symington LS. The dark side of homology-directed repair. DNA repair. 2021;106:103181.

85. Carvalho CM, Lupski JR. Mechanisms underlying structural variant formation in genomic disorders. Nat Rev Genet. 2016;17(4):224–38.

86. Wang WJ, Li LY, Cui JW. Chromosome structural variation in tumorigenesis: mechanisms of formation and carcinogenesis. Epigenetics Chromatin. 2020;13(1):49.

87. Littlefield C, Lazaro-Guevara JM, Stucki D, Lansford M, Pezzolesi MH, Taylor EJ, et al. A Draft Pacific Ancestry Pangenome Reference. bioRxiv. 2024:2024.08. 07.606392.

88. Nassir N, Almarri MA, Kumail M, Mohamed N, Balan B, Hanif S, et al. A draft Arab pangenome reference. bioRxiv. 2024:2024.07. 09.602638.

89. Olbrich M, Mousa M, Wohlers I, Aamri AA, Alnaqbi H, Alsuwaidi AH, et al. An Emirati pangenome incorporating a diploid telomere-to-telomere reference. bioRxiv. 2024:2024.12. 16.628631.

90. Collins RL, Talkowski ME. Diversity and consequences of structural variation in the human genome. Nature Reviews Genetics. 2025.

91. Eisfeldt J, Ameur A, Lenner F, Berk de Boer ET, Ek M, Wincent J, et al. Towards routine long-read sequencing for rare disease: a national pilot study on chromosomal rearrangements. medRxiv. 2023:2023.12. 15.23299892.

92. Geysens M, Huremagic B, Souche E, Breckpot J, Devriendt K, Peeters H, et al. Clinical evaluation of long-read sequencing-based episignature detection in developmental disorders. Genome Medicine. 2025;17(1):1.

93. Groza C, Schwendinger-Schreck C, Cheung WA, Farrow EG, Thiffault I, Lake J, et al. Pangenome graphs improve the analysis of structural variants in rare genetic diseases. Nature Communications. 2024;15(1):657.

94. Hiatt SM, Lawlor JM, Handley LH, Latner DR, Bonnstetter ZT, Finnila CR, et al. Long-read genome sequencing and variant reanalysis increase diagnostic yield in neurodevelopmental disorders. Genome Research. 2024;34(11):1747–62.

95. Marwaha S, Knowles JW, Ashley EA. A guide for the diagnosis of rare and undiagnosed disease: beyond the exome. Genome Medicine. 2022;14(1):23.

96. Negi S, Stenton SL, Berger SI, Canigiula P, McNulty B, Violich I, et al. Advancing long-read nanopore genome assembly and accurate variant calling for rare disease detection. The American Journal of Human Genetics. 2025;112(2):428–49.

97. Sinha S, Rabea F, Ramaswamy S, Chekroun I, El Naofal M, Jain R, et al. Long read sequencing enhances pathogenic and novel variation discovery in patients with rare diseases. Nature Communications. 2025;16(1):2500.

98. Steyaert W, Sagath L, Demidov G, Yépez VA, Esteve-Codina A, Gagneur J, et al. Unravelling undiagnosed rare disease cases by HiFi long-read genome sequencing. medRxiv. 2024.

99. Xu R, Zhang M, Tian W, Yang X, Li C. Decoding Complexity: The Role of Long-Read Sequencing in Unraveling Genetic Disease Etiologies. Mutation Research-Reviews in Mutation Research. 2025:108529.

100. Kubo Y, Nishio H, Saito K. A new method for SMN1 and hybrid SMN gene analysis in spinal muscular atrophy using long-range PCR followed by sequencing. J Hum Genet. 2015;60(5):233–9.

101. Woo EG, Tayebi N, Sidransky E. Next-Generation Sequencing Analysis of GBA1: The Challenge of Detecting Complex Recombinant Alleles. Front Genet. 2021;12:684067.

