## Supplementary Figures for "High-fidelity long-read sequencing reveals a complex RCCX locus at the single-nucleotide level in Korean patients with congenital adrenal hyperplasia"

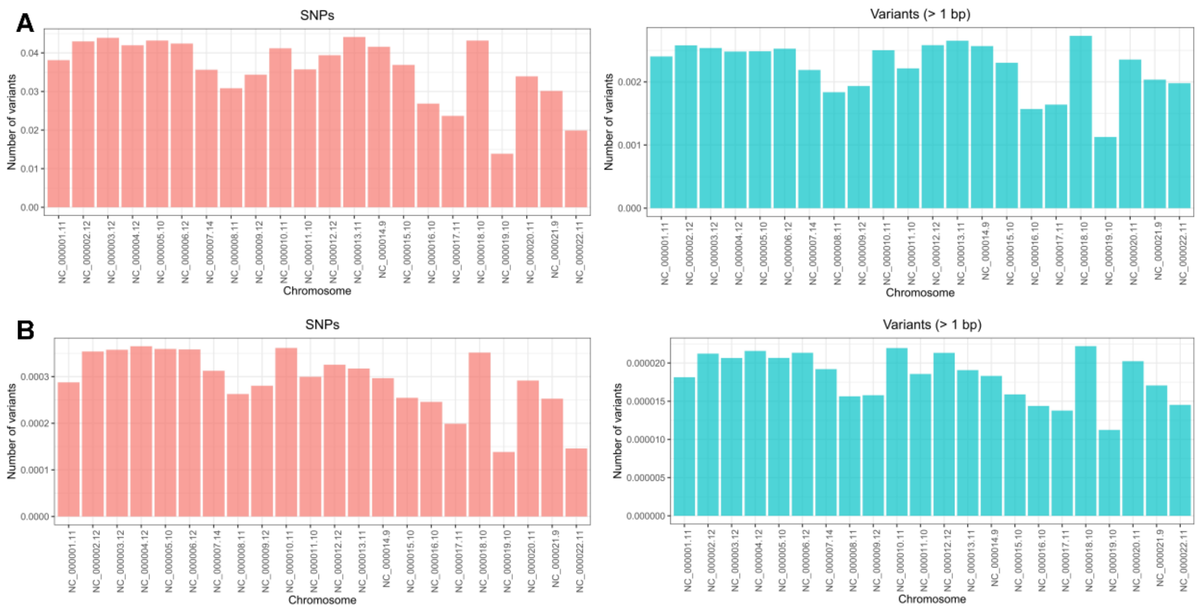


**Figure S1. Distribution of Korean-specific variant counts per chromosome.** The left panels show the distribution of SNPs, while the right panels display the distribution of variants greater than 1 bp. **A** represents the distribution of Korean-specific variants normalized by the total number of variants per chromosome, and **B** shows the distribution normalized by chromosome length.

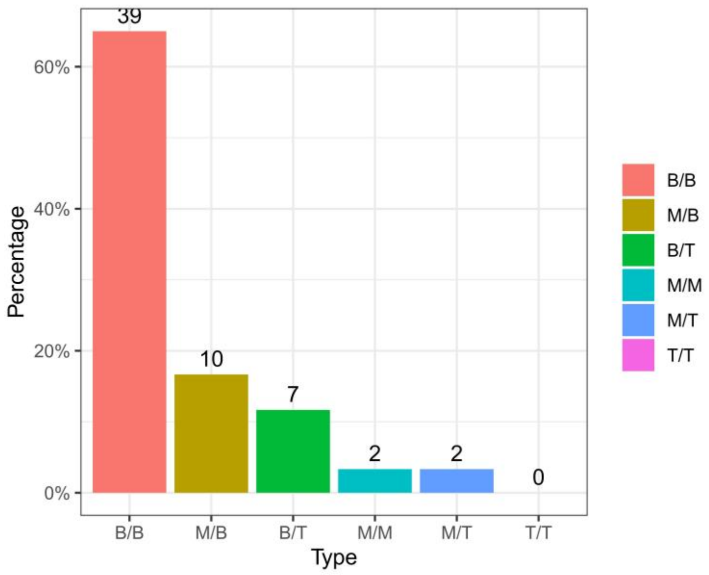


**Figure S2. Population-level zygosity analysis of RCCX modular structures.** The number of individuals corresponding to each bar is displayed on top of the bars. B, bimodular; M, monomodular; T, trimodular.
